# Clonal hematopoiesis and cell-free DNA characteristics in patients with high-risk large B-cell lymphomas

**DOI:** 10.64898/2026.10.14.26365378

**Authors:** Miro Nylén, Leo Meriranta, Emil Hiitola, Maare Arffman, Harald Holte, Judit Jørgensen, Peter Brown, Sirkku Jyrkkiö, Mats Jerkeman, Kristina Drott, Øystein Fluge, Magnus Björkholm, Lars Møller Pedersen, Mikael Eriksson, Satu Mustjoki, Sirpa Leppä, Mikko Myllymäki

## Abstract

Clonal hematopoiesis (CH) can predict treatment outcomes in solid cancers, but the prevalence and clinical impact of CH in previously untreated, aggressive large B-cell lymphomas (LBCL) remains poorly characterized. Moreover, CH may be detected in cell-free DNA (cfDNA)-based biomarker assays, but the longitudinal dynamics of CH mutations during and after immunochemotherapy in LBCL remain unknown. Hence, we characterized the spectrum of CH mutations in 247 patients with high-risk primary LBCL from three phase II clinical trials. We identified CH mutations in 23% of patients. Compared with healthy controls, *TET2* mutations were enriched in peripheral blood (PB) samples of LBCL patients (P=0.01). CH clone size showed high concordance between PB and cfDNA (R2=0.59, P<0.0001) but was not associated with circulating tumor DNA (ctDNA) concentration (P-values 0.25-0.86). In contrast with prior reports in the context of recurrent lymphoma, CH mutation status was not associated with overall survival (OS) (P=0.78) or progression-free survival (PFS) (P=0.89) in our primary LBCL cohort. CH clones showed variable longitudinal dynamics during immunochemotherapy, with 18% of evaluable patients developing mutant CH clones, mostly with *TP53*/*PPM1D* mutations.

Together, CH mutations can be readily detected in cfDNA of lymphoma patients, but they do not predict survival in primary high-risk LBCL.

## Introduction

The acquisition of somatic DNA variants in hematopoietic stem cells (HSCs) and their clonal expansion is a highly prevalent phenotype of aging [1], and these clonal hematopoiesis (CH) mutations contribute to shorter overall survival (OS), increased risk of hematologic malignancy, and various proinflammatory phenotypes at the population level [2–4]. The presence of CH mutations in the tumor microenvironment (TME) predicts improved immunotherapy responses [5, 6], but shorter disease-free survival in patients with solid tumors [7], highlighting the need for context-specific understanding of the clinical impact of CH in cancer patients.

The prevalence and the clinical impact of CH mutations in newly diagnosed aggressive non-Hodgkin lymphomas, including large B-cell lymphoma (LBCL), remain to be fully described. Among B-cell lymphomas, the incidence of CH mutations is most prevalent in indolent compared to aggressive B-NHL subtypes [8]. Moreover, mutation-carrying clones show a bias toward B-cell differentiation [8], in contrast to the myeloid-biased differentiation observed in aging HSCs carrying CH mutations in the general population [9]. To date, CH has mainly been studied in the context of relapsed and refractory (R/R) B-cell lymphomas. Specific CH mutations predict shorter survival and an increased risk of therapy-related myeloid neoplasia (TMN) after autologous stem cell transplantation in non-Hodgkin lymphomas [10, 11], whereas the association between CH mutations and chimeric antigen receptor (CAR) T cell therapy outcomes for B-cell lymphomas remains controversial [12–16]. Importantly, systematic evaluation of the impact of CH mutations in peripheral blood (PB) at diagnosis on long-term treatment outcomes has so far been limited by small cohort sizes, heterogeneous timing of DNA sampling, and particular focus on mantle cell lymphoma (MCL) [17–21].

Chemotherapy shapes the landscape of CH, particularly by expanding clones carrying *TP53* and *PPM1D* mutations [22–24]. Expansion of such clones with genetic DNA damage response (DDR) deficiency has been reported in peripheral blood and bone marrow samples of MCL patients during immunochemotherapy [19], and CH mutations present in R/R MCL may predispose patients to hematological toxicity and shorter survival [25]. However, the emergence and longitudinal dynamics of CH mutations in the peripheral blood of newly diagnosed high-risk LBCL treated with immunochemotherapy remain incompletely characterized.

Cell-free DNA (cfDNA) based detection of minimal residual disease (MRD) is emerging as a tool for disease monitoring in oncology [26, 27]. In LBCL, circulating tumor DNA (ctDNA) positivity after frontline therapy predicts disease recurrence [28–31] and may guide risk-adapted treatment approaches in the future [32]. In addition to tumor-derived extracellular DNA, a significant fraction of the mutated cfDNA molecules can originate from non-malignant hematopoietic cells [33], and these CH mutations may be misclassified as tumor-derived ctDNA [34, 35]. Therefore, characterizing the spectrum of CH mutations in cfDNA samples may have clinical implications, particularly in lymphomas, where CH clones may be clonally related to malignant cells in a subset of patients [8].

In the current study, we sequenced pretreatment peripheral blood samples from 247 primary high-risk LBCL patients enrolled in Nordic phase II clinical trials to detect CH mutations and evaluate their clinical impact. We traced the composition and cell fractions of CH mutations during and after therapy in the tumor, peripheral blood, and cfDNA compartments. Furthermore, we determined the associations between CH mutations, ctDNA features, and clinical outcomes and identified CH as a prevalent phenomenon in LBCL patients.

## Methods

### Nordic Lymphoma Group LBCL patient cohorts and healthy controls

We utilized three distinct cohorts of patients aged up to 65 years with primary high-risk LBCL who were treated in the Nordic Lymphoma Group (NLG) phase II trials with anthracycline-based dose-intensified immunochemotherapy and central nervous system (CNS) prophylaxis with high-dose methotrexate and high-dose cytarabine (LBC-04, -05, and -06) [32, 36, 37] (Table S1). We sequenced 38, 105, and 104 available whole blood samples from the LBC-04, LBC-05, and LBC-06 studies, respectively [32, 36, 37]. The median age was 55 years, 41% were female, and the median follow-up time of patients was 63.2 months. We also leveraged sequencing data of 163 healthy controls collected from the Finnish Red Cross Blood Service (Table S2) [4]. This study was conducted under the ethical approval number HUS/2215/2018 from the Ethics Committee of HUS (Helsinki, Finland). All participants provided informed consent to participate in one of the three phase II trials forming the basis for this study, and the principles of the Declaration of Helsinki were followed.

### Targeted next-generation sequencing (NGS) to detect clonal hematopoiesis in peripheral blood

We sequenced pretreatment (N=247) and follow-up (N=43) peripheral blood samples from LBCL patients using targeted sequencing of 65 myeloid driver genes (Table S3) as previously described [4]. Genomic DNA (50 ng) was processed according to Twist Library Preparation EF 2.0 with Enzymatic Fragmentation DOC-001239 REV 1.0 and Twist Target Enrichment Protocol DOC-001085 REV 2.0 manual (Twist Bioscience, San Francisco, CA, USA) with the following modifications. IDT xGEN unique dual index with unique molecular identifier adapters were used for ligation (Integrated DNA Technologies, Coralville, IA, USA). Library quantification and quality check was performed using LabChip GX Touch HT High Sensitivity assay (PerkinElmer, Waltham, MA, USA) and Qubit Broad Range DNA Assay (Thermo Fisher Scientific, Waltham, MA, USA). Libraries were pooled to 14-to 16-plex reactions. The target enrichment was performed using Twist custom panel probes (244 kb). The captured library pools were quantified for sequencing using QuantStudio5 Collibri Library Quantification kit (Thermo Fisher Scientific, Waltham, MA, USA) and LabChip GX Touch HT High Sensitivity assay (PerkinElmer, Waltham, MA, USA). Sequencing was performed with the Illumina NovaSeq6000 system (Illumina, San Diego, CA, USA) and v1.5 chemistry. The median target sequencing coverage was 1839x across samples.

We called CH variants from the targeted NGS panel sequencing data using Illumina DRAGEN 3.8 in tumor-only mode and used ANNOVAR for variant annotation. The DRAGEN commands are listed in Table S4. Only variants labeled PASS by DRAGEN were included for subsequent filtering. To filter benign and likely germline variation, we used a maximum population allele frequency threshold of 0.001 of any population in gnomAD (version 3.0), excluding known CH hotspots *JAK2* V617F and *DNMT3A* R882. We excluded synonymous and non-frameshift insertion/deletion variants. We also excluded variants with less than 300 total reads or with less than three variant allele reads. Furthermore, we required every variant to have at least one read in each direction supporting each allele. Only variants with a mosaic quality score of 150 or higher were included. We only included variants with a variant allele frequency (VAF) of at least 0.01. Finally, we manually curated detected variants by comprehensive examination of each variant individually in Integrative Genomics Viewer (version 2.19.7).

### Detection of CH variants in cell-free DNA and tumor DNA samples

We evaluated the identified CH mutations in pretreatment and follow-up plasma cfDNA samples of 104 patients of the LBC-06 cohort. The cfDNA samples were previously subjected to duplex sequencing using a targeted gene panel (Table S5). The lymphoma ctDNA results have been previously reported and were available for the current study [29, 32]. Besides encompassing lymphoma-related genomic regions, the panel covered the most common CH driver genes for the purposes of the current work (Table S5) [25]. In addition to cfDNA samples, the same gene panel was applied to available diagnostic tumor DNA samples, including eleven from patients with CH mutations detected in PB. VAFs of the CH mutations identified in PB with the myeloid panel were inferred from the cfDNA, tumor DNA, and PB sequences as previously described [32], without discriminating between duplex and simplex reads and setting mapping and base quality at 30.

## Statistical analysis

Genotypes were assigned before evaluating the association with clinical variables, including outcomes. Analyses were performed using R and GraphPad Prism. Linear regression and logistic regression models were fitted using the R package stats (version 4.5.2), and GraphPad Prism (version 10.6.1), survival (version 3.8.5) was used for Cox proportional hazards (Cox-PH) models, and Kaplan-Meier models were fitted using GraphPad Prism (version 10.6.1). Stats (version 4.5.2) and CATT (version 2.0) were used for statistical tests. P-values reported are two-sided. P-values < 0.05 were considered statistically significant throughout the study.

Multivariable models were adjusted for age and sex in prevalence comparisons, and for age, sex and age-adjusted International Prognostic Index (aaIPI) in survival analyses.

## Results

### CH mutations in newly diagnosed aggressive LBCL

To evaluate the prevalence and spectrum of CH mutations in pretreatment PB samples of newly diagnosed patients with high-risk LBCL, we performed targeted sequencing of recurrently mutated CH genes (Table S3) [4] in 247 LBCL patients treated in NLG-LBC phase II trials (Table S1) [32, 36, 37]. We then integrated CH information with pretreatment cfDNA characteristics, clinical information, and sequencing data from longitudinal PB and cfDNA samples to elucidate the clinical impact of CH in LBCL (Fig. 1A). Additionally, we leveraged sequencing data of 163 healthy blood donors (Table S2) [4]. The median age at inclusion was 55 years, and patients had clinically high-risk disease (aaIPI 2-3) and/or risk factors for CNS recurrence. Treatment consisted of anthracycline-based intensified immunochemotherapy and CNS prophylaxis. The combined 3-year OS and progression-free survival (PFS) rates were 88% and 81%, respectively.

**Figure 1.**
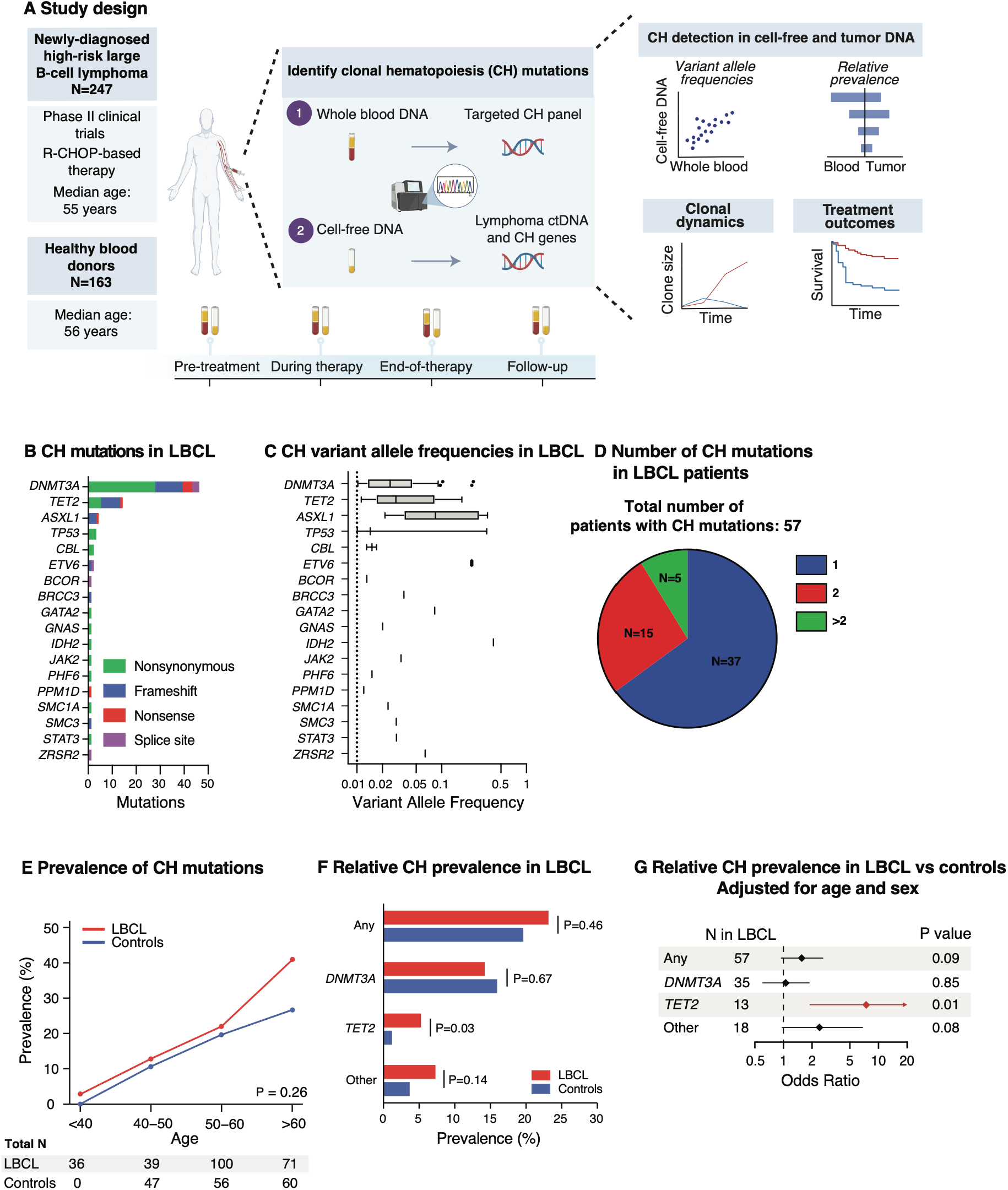
Clonal hematopoiesis mutations in newly diagnosed high-risk B-cell lymphoma. (A) Outline of the study. (B) Distribution of CH mutations by gene in LBCL patients, further annotated by variant type. (C) Distribution of VAFs by mutated gene. (D) Total number of mutations in LBCL patients. (E) Prevalence of CH by age brackets in LBCL patients and healthy controls. The difference in trend between groups was assessed using logistic regression. (F) Comparison of CH prevalences in LBCL versus healthy controls. P-values were calculated using Fisher’s exact test. (G) Multivariable logistic regression models for CH prevalence adjusted for age and sex.

We identified a total of 83 CH mutations with over one percent VAF in pretreatment PB samples from 57 out of 247 (23%) of patients (Fig. 1B, Table S6, methods). In contrast to R/R B-cell lymphoma [10, 11], the classical aging-related CH genes *DNMT3A*, *TET2*, and *ASXL1* [1] showed the highest number of variants in our cohort (Fig. 1B). Variants in *IDH2* (median VAF, 0.41) and *ETV6* (median VAF, 0.23) genes had the highest median cell fractions in PB (Fig. 1C). The majority (37/57, 65%) of patients harbored a single CH mutation, while 20 patients (35%) presented with multiple co-occurring CH alterations (Fig. 1D).

The median VAF of CH mutations in PB was similar between LBCL patients and healthy blood donors (median VAF, 0.026 vs 0.023, P=0.30). The overall prevalence of CH was not significantly different between LBCL and controls (prevalence, 23% vs 20%, P=0.46) (Fig. 1E-G). However, variants in *TET2* were significantly enriched in LBCL patients compared with healthy controls (P=0.03) (Fig. 1F, Fig. S1), and this difference remained significant in a multivariable model adjusted for age and sex (P=0.01) (Fig. 1G). This observation is consistent with the putative role of *TET2* mutations in driving B-cell lymphoma pathogenesis [38–40]. Mutations in other genes showed similar prevalences between LBCL and controls (Fig. 1F-G).

### Clinical and molecular correlates of CH mutations in LBCL

Next, we evaluated patient characteristics by CH status at the time of LBCL diagnosis (Table 1, Fig. 2A). The prevalence of CH mutations was 23%, ranging from 11% to 27% across the three trials (Table S7). As expected, CH carriers were older than patients without CH mutations (median age, 60 years versus 54 years, P<0.001). We did not observe any differences between CH status and ECOG performance score (P=0.29), Ann Arbor disease stage (P=0.12), age-adjusted IPI score (P=0.16), bone marrow infiltration (P=0.82), B symptoms (P>0.99), or elevated lactate dehydrogenase (LDH) levels (P=0.18) (Fig. 2A, Table 1).

**Figure 2.**
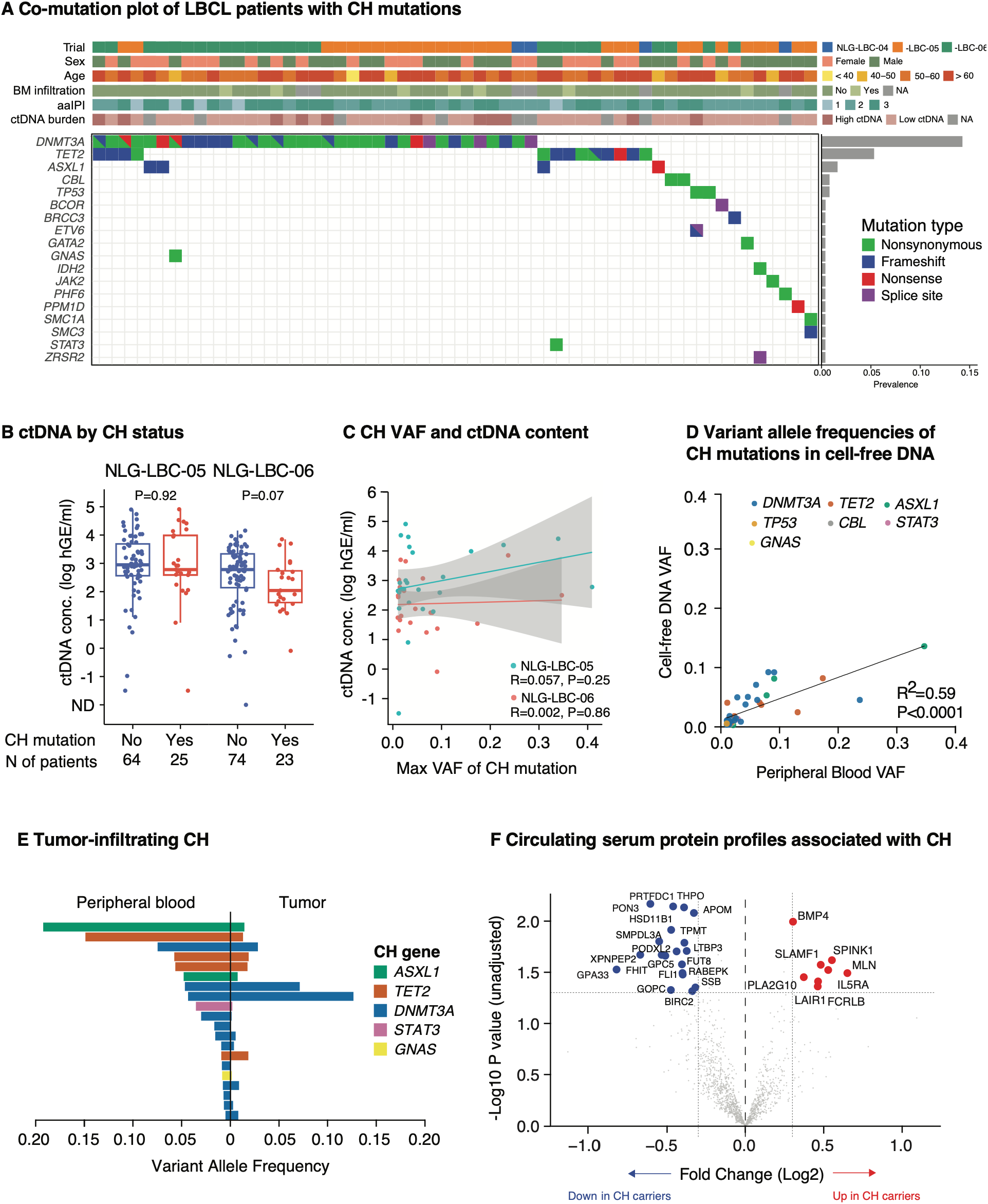
Detection of clonal hematopoiesis in cell-free DNA. (A) Patient-level CH mutation landscapes and their association with clinical features. (B) Total ctDNA burden by CH status in the LBC-05 and LBC-06 clinical cohorts by CH mutation status. The difference within each cohort was assessed using the Wilcoxon rank-sum test. (C) The association between maximum CH VAF and total ctDNA burden in the LBC-05 and LBC-06 cohorts. R^2^ and P-value were calculated using linear regression. (D) Corresponding VAFs of CH mutations in PB and cfDNA. R^2^ and P-value were calculated using linear regression. (E) The corresponding VAFs of CH mutations in PB and tumor DNA. (F) Serum protein profiles by CH mutation status. NPX data were generated by Olink assays and nominal P values were calculated using empirical Bayes moderated t-test.

**Table 1.**
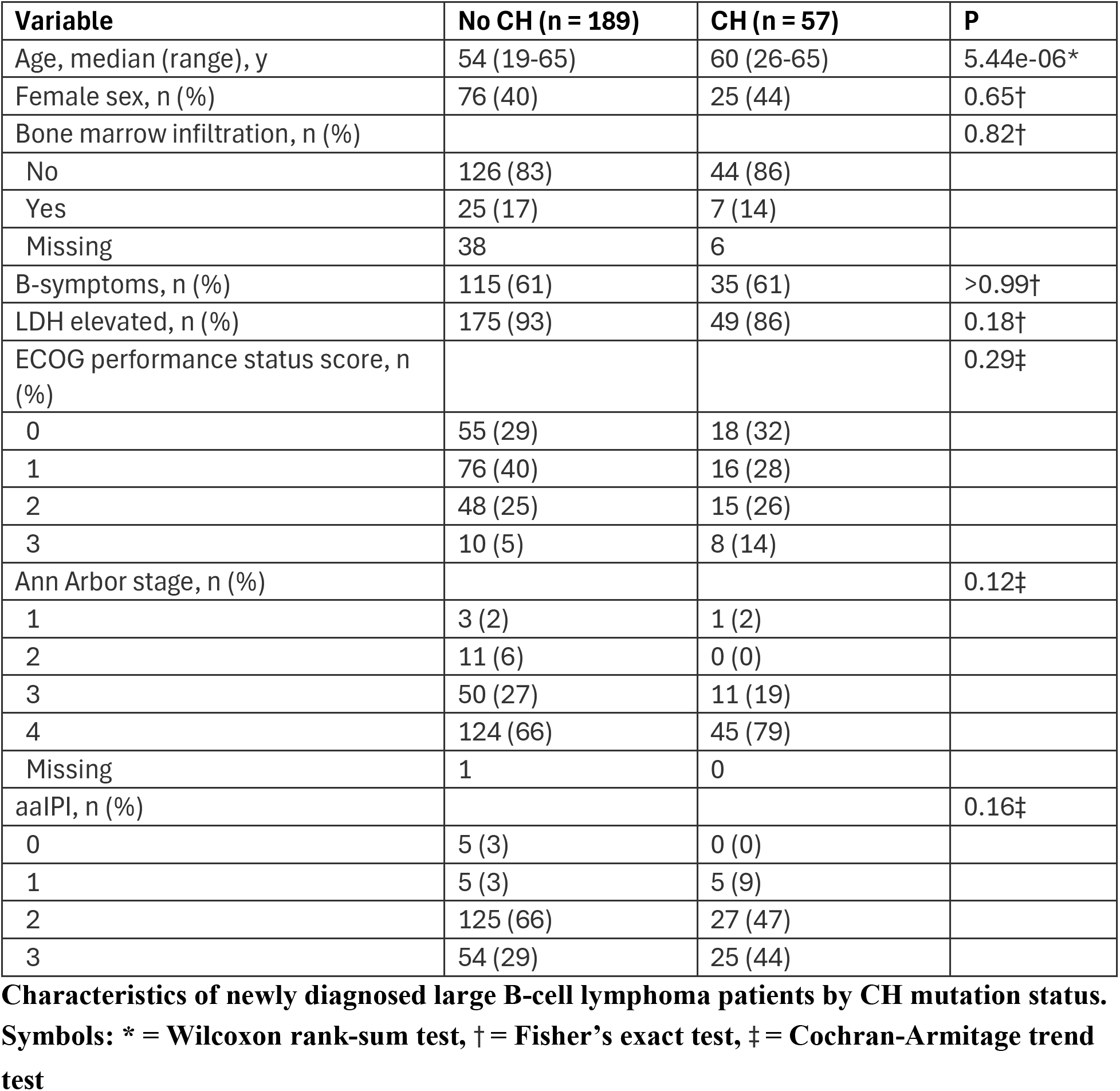
Patient characteristics of newly diagnosed large B-cell lymphoma patients.

| **Variable** | **No CH (n = 189)** | **CH (n = 57)** | **P** |
| --- | --- | --- | --- |
| Age, median (range), y | 54 (19-65) | 60 (26-65) | 5.44e-06* |
| Female sex, n (%) | 76 (40) | 25 (44) | 0.65† |
| Bone marrow infiltration, n (%) |  |  | 0.82† |
| No | 126 (83) | 44 (86) |  |
| Yes | 25 (17) | 7 (14) |  |
| Missing | 38 | 6 |  |
| B-symptoms, n (%) | 115 (61) | 35 (61) | >0.99† |
| LDH elevated, n (%) | 175 (93) | 49 (86) | 0.18† |
| ECOG performance status score, n (%) |  |  | 0.29‡ |
| 0 | 55 (29) | 18 (32) |  |
| 1 | 76 (40) | 16 (28) |  |
| 2 | 48 (25) | 15 (26) |  |
| 3 | 10 (5) | 8 (14) |  |
| Ann Arbor stage, n (%) |  |  | 0.12‡ |
| 1 | 3 (2) | 1 (2) |  |
| 2 | 11 (6) | 0 (0) |  |
| 3 | 50 (27) | 11 (19) |  |
| 4 | 124 (66) | 45 (79) |  |
| Missing | 1 | 0 |  |
| aalPI, n (%) |  |  | 0.16‡ |
| 0 | 5 (3) | 0 (0) |  |
| 1 | 5 (3) | 5 (9) |  |
| 2 | 125 (66) | 27 (47) |  |
| 3 | 54 (29) | 25 (44) |  |

**Characteristics of newly diagnosed large B-cell lymphoma patients by CH mutation status.**
**Symbols: \* = Wilcoxon rank-sum test, † = Fisher's exact test, ‡ = Cochran-Armitage trend test**

The pretreatment ctDNA concentration was available for 186 patients (Table S1) and was not higher among CH mutation carriers (Fig. 2B). Furthermore, CH clone size was not associated with ctDNA concentration (Fig. 2C), indicating that ctDNA levels are independent of CH mutation status in newly diagnosed LBCL.

CH mutations detected in PB show similar representation in cfDNA among individuals without cancer [41], but the relative proportion of CH clone-derived DNA molecules in cfDNA of LBCL patients is unknown. For CH mutations present in PB at the time of diagnosis, we examined their relative abundance in concurrent plasma cfDNA (methods). All 40 CH mutations detected in PB of patients with evaluable concurrent pretreatment cfDNA were also detected in plasma cfDNA. Overall, the CH mutation VAFs in cfDNA were associated with PB VAFs (R^2^= 0.59, P-value<0.001, Fig. 2D).

The presence of CH mutations in tumor tissue may influence treatment outcomes among patients with solid tumors in a context-dependent manner [5–7]. Therefore, we analyzed the association of CH mutation VAFs across tumor, blood, and cfDNA samples in patients for whom tumor genotypes were available (Fig. S2, Table S1). Eleven of the 24 CH patients with a total of 20 CH mutations had available tumor DNA sequenced. We detected 17 of the 20 CH mutations (85%) in the corresponding tumor samples from 9 of the 11 patients (82%) with a mean VAF of 0.020 (range 0.0006–0.126) (Table S8). Eight of the 20 CH mutations (40%) were present in the tumor with at least one percent VAF in 5 of the 11 CH patients (Fig. 2E). Moreover, PB VAFs did not associate with tumor VAFs (R^2^= 2.1e-2, *P* = 0.54), and a similar lack of association was observed for the VAFs from plasma cfDNA and tumor (R^2^= 8.6e-2, *P* = 0.21, Fig. S2). Together, these findings indicate that while CH mutations are prevalent in pretreatment plasma cfDNA in LBCL, tumor-infiltrating CH (TI-CH) is less prevalent [42].

We explored the association between CH mutation status and previously reported serum protein profiles from 94 patients (Table S1) [43]. We observed higher levels of eight and lower levels of 19 serum proteins in patients with LBCL (Fig. 2F); however, none of these associations remained significant after correcting for multiple hypothesis testing. Similar observations were made for patients with *DNMT3A* and *TET2* mutations (Fig. S3A-D). Notably, CH mutations were not enriched in patients with a previously described inflamed serum protein profile (Fig. S3E) [43]. In addition, focused analyses of the cytokines previously linked with CH mutations in the population level [44] revealed no significant differences with respect to CH status in LBCL (Fig. S3F).

### Pretreatment CH and frontline treatment outcomes

To assess the prognostic value of CH mutations at the time of diagnosis, we performed univariable and multivariable models for OS and PFS. CH mutations were not associated with OS in a univariable (P=0.78, Fig. 3A) or in a multivariable (P=0.61, Fig. 3C) analysis adjusting for age, sex, and aaIPI. This finding was further replicated in gene-specific univariable models (Fig. S4) and across trials (Fig. S5). Similarly, CH mutations showed no association with PFS in univariable (P=0.89, Fig. 3B; Fig. S4) or multivariable (P=0.74, Fig. 3D; Fig. S5) models. While potentially limited by statistical power to detect gene-specific effects, our results indicate that CH variants present in pretreatment samples do not contribute to the outcomes of patients aged up to 65 years with clinically high-risk LBCL.

**Figure 3.**
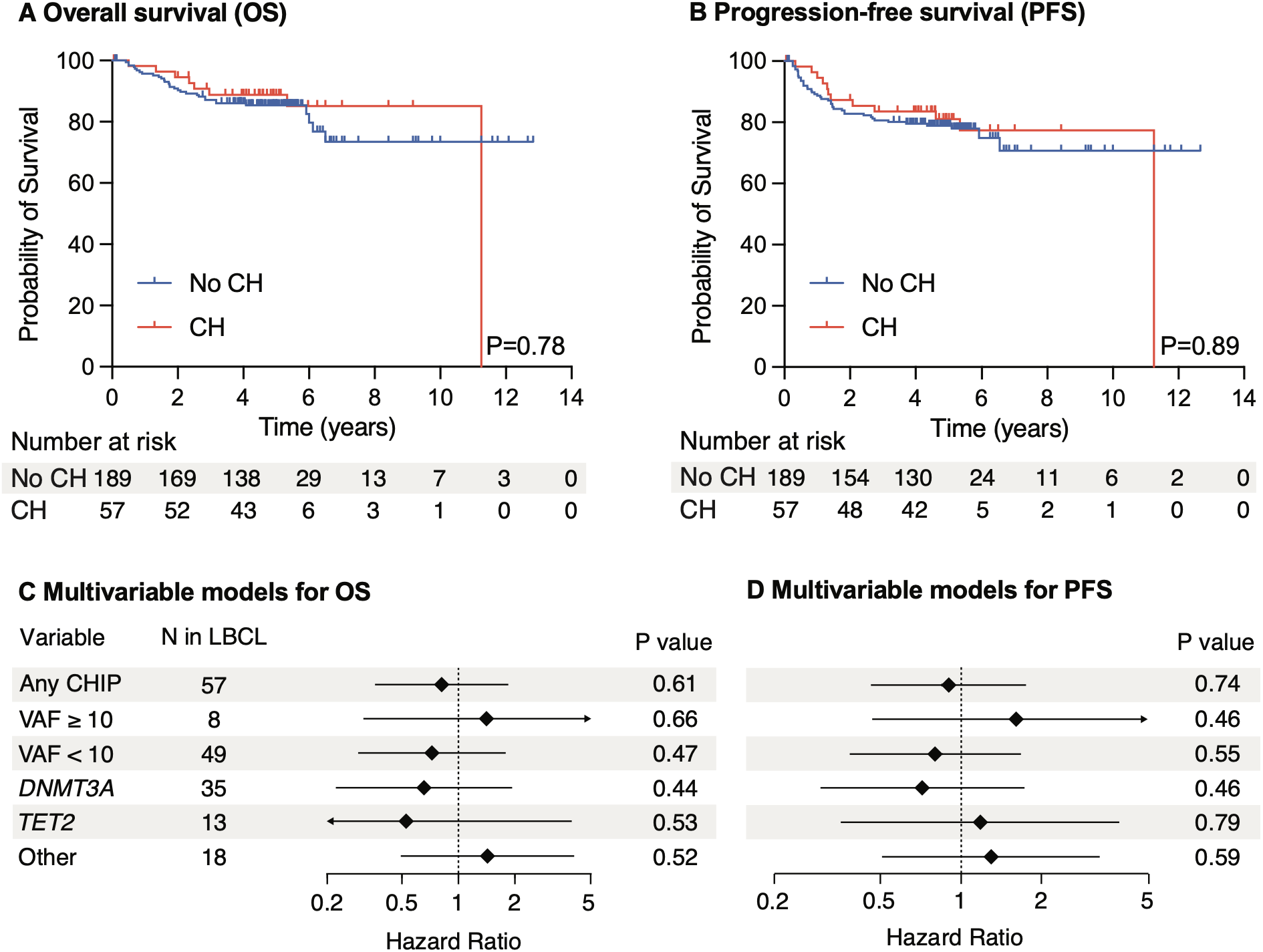
The impact of CH mutations on patient outcomes. (A) Kaplan-Meier for OS by CH mutation status. (B) Kaplan-Meier for PFS by CH mutation status. (C) Cox PH models for OS by CH mutation status. (D) Cox PH models for PFS by CH mutation status. (C) and (D) are adjusted for age, sex, and aaIPI.

### Longitudinal dynamics and emergence of CH variants in LBCL

Antineoplastic therapy may shape expansion rates of CH clones [10, 23], with potential long-term implications for clinical outcomes such as the risk of therapy-related myeloid malignancy. To characterize the clonal dynamics of pre-existing CH mutations during high-dose immunochemotherapy in LBCL, we sequenced serial PB samples and analyzed them alongside previously sequenced serial cfDNA samples (Table S1). CH mutations showed variable changes in PB VAFs between the two time points (Fig. 4A). While *DNMT3A* and *TET2* mutations showed stable VAFs during follow-up (median VAF difference, 0.004 and -0.003, respectively, Fig. 4A), two mutations in *SMC1A/3* genes showed clonal expansion during therapy (Fig. 4A-B). Out of the 43 LBCL patients with available follow-up PB samples, 6 (14%) showed the emergence of new *TP53*/*PPM1D* clones (median VAF, 0.016) following therapy (Fig. 4C), consistent with the high prevalence of *TP53* and *PPM1D* mutations in pretreated patients [10].

**Figure 4.**
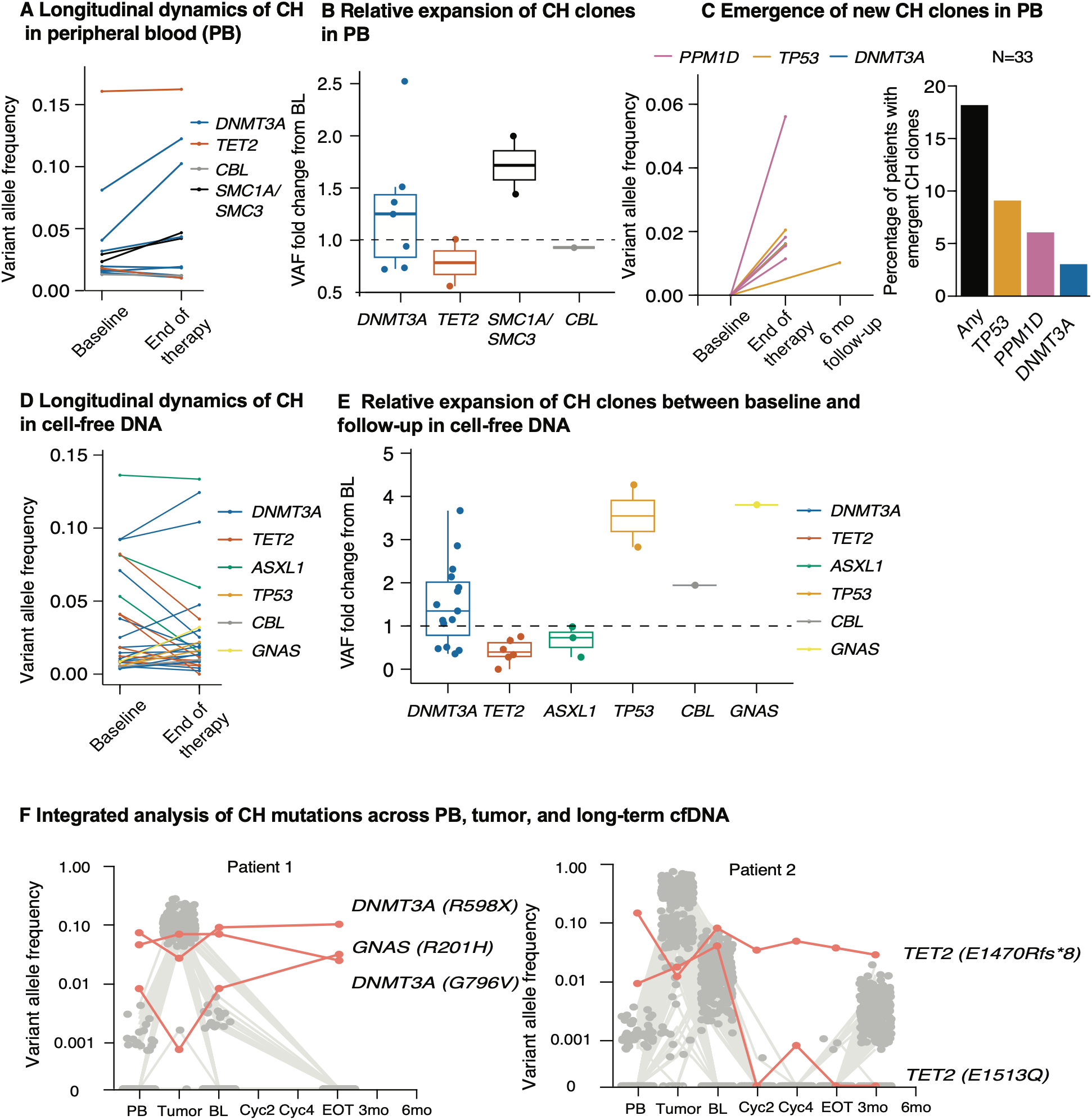
Longitudinal dynamics and emergence of CH variants in LBCL. (A) VAF dynamics of CH mutations during follow-up PB. (B) Relative expansion of CH clones in PB from baseline to end of therapy. (C) Emergence of new CH mutations in PB during follow-up. (D) VAF dynamics of CH mutations during follow-up in cfDNA. (E) Relative expansion of CH clones in cfDNA by the end of therapy compared to baseline. (F) VAF dynamics of CH variants and lymphoma reporter mutations across PB, tumor, and cfDNA samples at pretreatment and in serial cfDNA samples during and after therapy.

Among the 33 patients without any CH at baseline, 18% developed new CH mutant clones on therapy (Fig. 4C). In the LBC-06 cohort, 123 patients were previously profiled for longitudinal plasma ctDNA using a targeted gene panel that included CH genes, enabling their profiling in the study context. 18 (75%) of patients with CH in cfDNA at baseline also had an end-of-therapy plasma sample available for longitudinal CH variant detection. After therapy, *TP53* mutant CH clones showed marked expansion from baseline (median fold change, 3.55), along with expansion of single mutations in *GNAS* (fold change, 3.81) and *CBL* (fold change, 1.94) genes (Fig. 4E). In contrast, *TET2* variants showed a relative decrease in clone size compared to pretreatment samples (median fold change, 0.40). *DNMT3A* (median fold change, 1.35) and *ASXL1* (median fold change, 0.73) clones remained largely stable over the treatment course (Fig. 4E).

The longitudinal dynamics of CH variants in cfDNA strongly contrasted with those of lymphoma reporter mutations, as most of the CH variants, 27 of the total 28 (96%), were still detectable in plasma cfDNA at the EOT (median VAF, 0.016; Fig. S6). The VAFs of most CH variants remained stable in plasma over the follow-up period (Fig. S6). For example, two *DNMT3A* and one *GNAS* variant in patient 1 showed variable VAFs (0.07, 0.05, 0.008, respectively) in PB at diagnosis, with tumor VAFs lower but cfDNA VAFs higher than those for lymphoma variants (Fig. 4F). However, the VAFs remained stable at the EOT despite clearance of all lymphoma variants in cfDNA (Fig. 4F). Therefore, the three mutations likely represented CH clones clonally distinct from the lymphoma clone that were also present in the TME at diagnosis. In representative case 2, we detected two *TET2* variants with VAFs of 0.009 and 0.15 in PB at diagnosis. Interestingly, despite similar representation in tumor and cfDNA at diagnosis, the two *TET2* clones showed very distinct clonal dynamics in longitudinal cfDNA samples. Variant E1470Rfs*8 showed persistent VAFs in the EOT and other follow-up samples despite clearance of the lymphoma clone in ctDNA, consistent with the likely non-lymphoma origin of the variant. In contrast, the other *TET2* variant (E1513Q) became undetectable at EOT and showed no expansion at disease relapse 3 months later (Fig. 4F). Therefore, the relative prevalence of CH mutations in cfDNA and tumor DNA at diagnosis is insufficient in predicting clonal origins and longitudinal dynamics of CH mutations during therapy.

We explored whether cfDNA characteristics may help distinguish lymphoma-driving *TET2* mutations from aging-related clonal hematopoiesis in LBCL. In 5 out of 7 patients, the VAF of *TET2* CH mutations in cfDNA was lower than the median VAF of lymphoma variants (Fig. S7). While these analyses cannot distinguish subclonal *TET2* lymphoma variants from clonally unrelated CH, the data suggest that the lymphoma clones were unlikely to originate from a non-malignant *TET2* CH clone in these patients. In further support of this, *TET2* mutations did not consistently show high VAFs in tumor DNA (Fig. 2E).

## Discussion

The prevalence and clinical impact of CH mutations in high-risk aggressive B-cell lymphomas have remained elusive. In this study, we leverage samples from three Nordic phase II first-line clinical trials and provide a comprehensive characterization of CH mutations at the time of LBCL diagnosis. Integrated analysis of PB, cfDNA, and tumor DNA sequencing shows variable representation of CH mutations across sample types. Furthermore, sequencing of longitudinal blood and DNA samples provides insights into the clonal dynamics of CH during high intensity immunochemotherapy and after completion of frontline treatment. To our knowledge, this is one of the first large-scale studies in LBCL in which CH status was uniformly assessed at diagnosis and its clinical implications were evaluated in treatment-naïve patients.

The overall prevalence of CH mutations at diagnosis using a 0.01 VAF threshold was 23%. While the prevalence of CH mutations was similar between LBCL patients and healthy controls, *TET2* mutations were enriched in LBCL. While we are unable to confidently determine the clonal relationship between CH mutations and lymphoma clones, our results suggest heterogeneous associations between the two. For example, our index case with two co-occurring *TET2* mutations showed markedly distinct clonal dynamics during lymphoma remission despite similar cell fractions in tumor and cfDNA at diagnosis (Fig. 4F), highlighting the heterogeneous behavior of CH clones over the course of lymphoma-targeting primary therapy. A recent study by Wiegand et al [8] used flow cytometry-based sorting of blood cell compartments from 43 patients with non-Hodgkin B-cell lymphoma and showed multilineage propagation of CH mutations in these patients, suggesting that CH clones can differentiate into lymphoid lineage cells in malignant B-cell lymphomas.

CH mutations were readily detectable in plasma cfDNA and showed high concordance of allele frequencies between PB and cfDNA, despite significant enrichment of tumor-derived ctDNA in these samples. One study showed a similar association between PB and cfDNA cell fractions in the non-malignant context [41]. As ctDNA measurement [30] as well as CH detection [7] may have predictive and prognostic effects in solid cancers, our results suggest that both compartments can be readily detected in single cfDNA samples with novel assays.

Fragmentomic analyses [25, 45] and machine learning-based algorithms [46] of cfDNA may help differentiate tumor-derived versus CH-derived origins of mutations, with potential clinical implications in the future.

In contrast with prior studies in previously treated B-cell lymphomas, we observed no association between CH mutation status and outcomes in our untreated patient population. The trial populations consisted of relatively young patients (median age, 55 years) with aggressive LBCL disease. Therefore, we acknowledge the potential for limited generalizability to older patients with higher CH prevalence [21]. Likewise, the gene-specific analyses are limited by the number of mutation carriers, requiring validation in larger studies. Prior literature evaluating the prognostic impact of CH in LBCL is mostly limited to the context of ASCT, where the clinical impact has been considered alongside other NHL subtypes [10, 11]. On the other hand, we noted expansion and emergence of CH clones carrying *TP53* and *PPM1D* mutations, consistent with similar findings in non-LBCL clinical contexts [10, 23]. Therefore, further studies with extended follow-up are needed to determine whether CH expansion influences long-term outcomes, such as the risk of secondary malignancy or cardiovascular disease risks in LBCL patients. Moreover, a recent study by Chen et al [42] showed an association between TI-CH and LBCL outcomes, warranting deeper understanding of the biological and clinical significance of the interaction between blood CH and TI-CH.

In summary, we show a high prevalence of CH mutations in previously untreated, high-risk LBCL patients. CH mutations also showed concordant representation in cfDNA, with variable allele frequencies in LBCL tumor DNA. While CH mutations were not associated with treatment outcomes, CH clones expanded in PB and cfDNA during immunochemotherapy. These observations warrant studies evaluating the impact of CH on long-term outcomes, such as the risk of therapy-related myeloid malignancies, in LBCL.

## Data availability

In accordance with European Union legislation, the General Data Protection Regulation (GDPR) and the Finnish Act on the Secondary Use of Health and Social Data, the clinical annotations and the linked sequencing data from this study cannot be deposited or shared publicly due to the sensitive patient data protection. Collaboration agreements, compliant with the above and accepted by the local ethics committees and authorities, can be queried from the last authors.

Processed sequencing data supporting the findings of this study are available in the supplementary materials.

## Data Availability

In accordance with European Union legislation, the General Data Protection Regulation (GDPR) and the Finnish Act on the Secondary Use of Health and Social Data, the clinical annotations and the linked sequencing data from this study cannot be deposited or shared publicly due to the sensitive patient data protection. Collaboration agreements, compliant with the above and accepted by the local ethics committees and authorities, can be queried from the last authors. Processed sequencing data supporting the findings of this study are available in the supplementary materials.

## Acknowledgements

The authors would like to thank HiPREP Core at FIMM Technology Centre, supported by HiLIFE and Biocenter Finland for DNA extraction services. Library preparation, sequencing, and DRAGEN analysis of LBCL patient and control samples were performed by FIMM Genomics NGS Sequencing Unit at the University of Helsinki, supported by HiLIFE and Biocenter Finland. The Center for Science Ltd. (CSC) is acknowledged for data storage and computational resources. We acknowledge the Finnish Red Cross Blood Service for providing healthy control samples.

## Author contributions

Conceptualization: M.N., L.M., S.L., and M.M. Methodology: M.N., L.M., S.L., and M.M. Software: E.H. Validation: M.N., L.M., and M.M. Formal analysis: M.N., L.M., E.H., M.A., and M.M. Resources: H.H., J.J., P.B., S.J., M.J., K.D., O.F., M.B., L.M.P., M.E., S.M., S.L., and M.M. Data curation: M.N., L.M., E.H., M.A., S.L., and M.M. Visualization: M.N, L.M., E.H., and M.M. Supervision: M.M., S.M., and S.L. Writing—original draft: M.N., L.M., and M.M. Writing—review and editing: All. Project administration: S.L. and M.M. Funding acquisition: S.M., S.L., and M.M.

## Ethics statement

This study was conducted under the ethical approval number HUS/2215/2018 from the Ethics Committee of HUS (Helsinki, Finland). All participants provided informed consent to participate in one of the three phase II trials forming the basis for this study, and the principles of the Declaration of Helsinki were followed.

