## Supplemental Figures and Table Legends for "Clonal hematopoiesis and cell-free DNA characteristics in patients with high-risk large B-cell lymphomas"

**Supplemental figure legends**

**Supplemental table legends**

**Supplemental figures**

### Supplemental figure legends

#### Figure S1. Distribution of TET2 mutations in LBCL and healthy controls

Distribution of *TET2* mutations in LBCL patients and healthy controls, annotated by variant type and shown in relation to the position of *TET2* functional domains.

#### Figure S2. Association between PB, cfDNA, and tumor variant allele frequencies of CH mutations

(A) Corresponding VAFs of CH mutations in peripheral blood and tumor. (B) Corresponding VAFs of CH mutations in cell-free DNA and tumor. $R^2$ and P-values were calculated using linear regression.

#### Figure S3. Differentially expressed proteins by CH mutation status

(A) Serum protein profiles by overall CH mutation status. (B) Serum protein profiles by *DNMT3A* mutation status. (C) Serum protein profiles by *TET2* mutation status. (D) Serum protein profiles by CH mutation status for genes other than *DNMT3A* or *TET2*. (E) The association between CH mutation status and the inflamed serum protein profile previously described by Arffman et al (PMID: 38579729). (F) The association between CH status and cytokines previously linked with CH mutations at the population level by Bick et al (PMID: 33057201). NPX data were generated by Olink assays and nominal P values were calculated using empirical Bayes moderated t-test.

#### Figure S4. Overall survival (OS) and progression-free survival (PFS) by mutated CH gene

(A) Kaplan-Meier for OS and PFS by *DNMT3A* mutation status. (B) Kaplan-Meier for OS and PFS by *TET2* mutation status. (C) Kaplan-Meier for OS and PFS by *ASXL1* mutation status. (D) Kaplan-Meier for OS and PFS by CH mutation status for genes other than *DNMT3A*, *TET2*, or *ASXL1*.

#### Figure S5. Multivariable models for OS and PFS in each trial

(A) Cox PH models for OS and PFS by CH mutation status in NLG-LBC-04 patients. (B) Cox PH models for OS and PFS by CH mutation status in NLG-LBC-05 patients. (C) Cox PH models for OS and PFS by CH mutation status in NLG-LBC-06 patients.

**Figure S6. CH variant allele frequencies in PB, tumor, and cfDNA**

VAF dynamics of CH mutations and lymphoma reporter mutations across PB, tumor, and cfDNA samples at pretreatment, as well as in serial cfDNA samples collected during and after therapy.

**Figure S7. Relative variant allele frequencies of TET2 and lymphoma variants in cfDNA**

Variant allele frequencies of *TET2* CH variants and lymphoma reporter mutations in cell-free DNA at pretreatment relative to corresponding PB VAFs for *TET2* CH variants.

### Supplemental table legends

**Table S1. NLG-LBC trials included in the study**

**Table S2. Baseline characteristics of healthy blood donors**

**Table S3. CH genes covered by the PB sequencing panel**

**Table S4. DRAGEN commands**

**Table S5. Targeted genomic regions (hg38) of the cfDNA sequencing panel with 10 base pair paddings**

**Table S6. CH mutations detected in newly diagnosed LBCL cohorts and healthy controls**

**Table S7. CH mutation prevalence in NLG-LBC trials**

**Table S8. Variant allele frequencies of clonal hematopoiesis (CH) mutations in different compartments and follow-up cell-free DNA. PB, peripheral blood; BL, pretreatment plasma; CYC, after cycle; tumor, diagnostic tumor DNA; EOT, at end-of-therapy; FU, follow-up month**

**Figure S1. Distribution of TET2 mutations in LBCL and healthy controls**

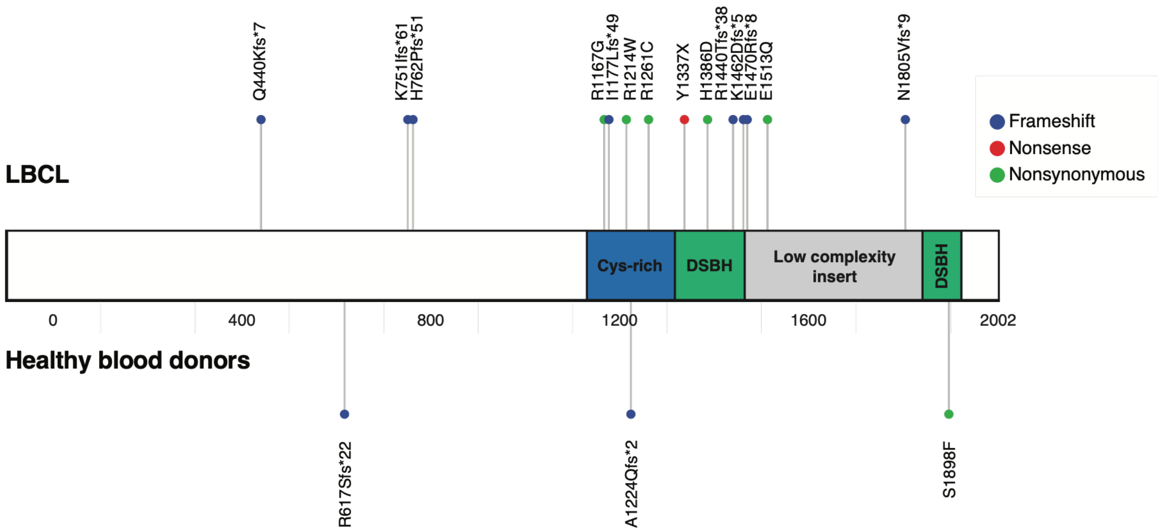

**Figure S2. Association between PB, cfDNA, and tumor variant allele frequencies of CH mutations**

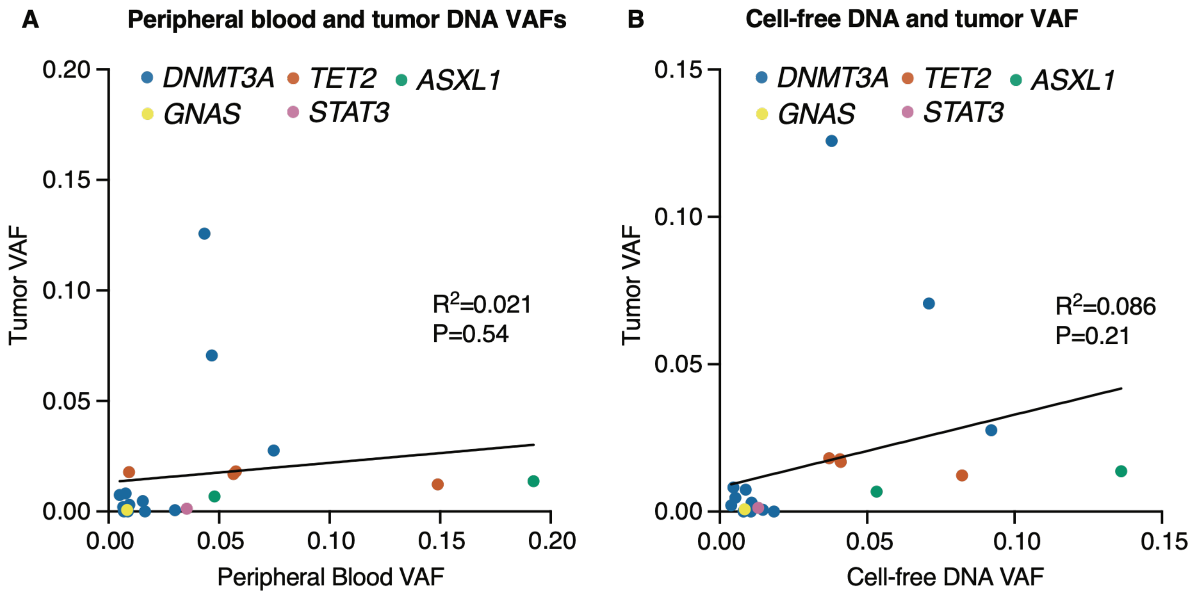

### Figure S3. Differentially expressed proteins by CH mutation status

#### A Any CH mutation

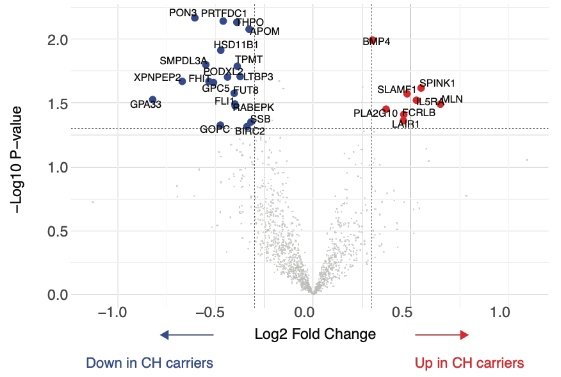


#### B DNMT3A

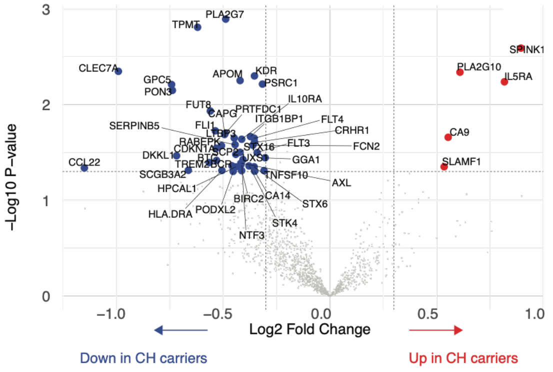


#### C TET2

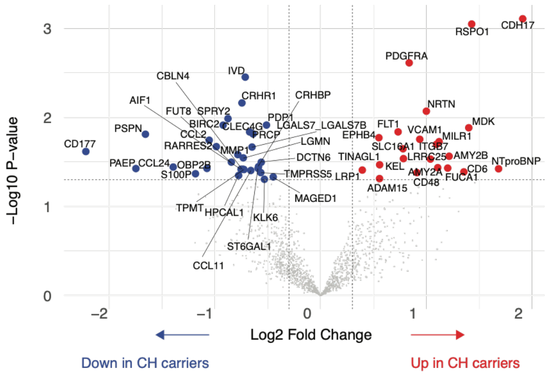


#### D Other CH mutations

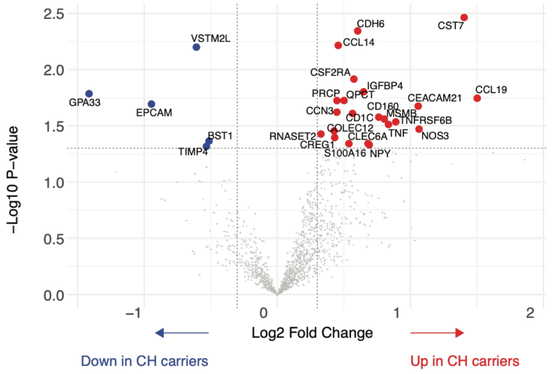


#### E CH mutation status and serum inflammatory profile

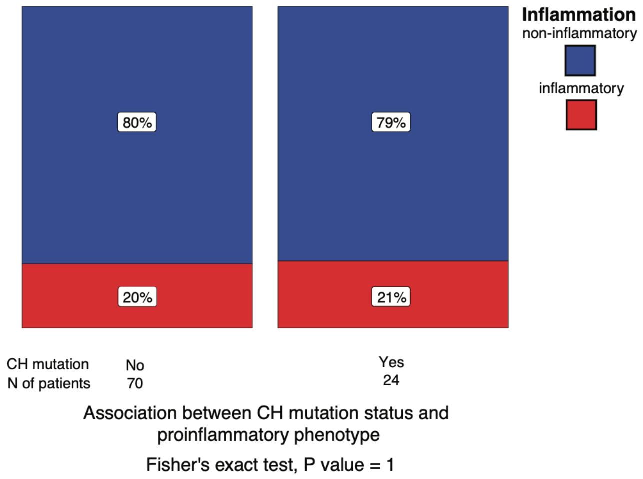


#### F Adjusted P values of CH-associated inflammatory cytokines

| | ALL | DNMT3A | Other | TET2 |
|---|---|---|---|---|
| IL18 | $Log_2FC$=0.12, P=0.99 | $Log_2FC$=-0.18, P=0.99 | $log_2FC$=0.54, P=1.0 | $Log_2FC$=0.61, P=0.98 |
| IL6 | $Log_2FC$=0.28, P=0.99 | $Log_2FC$=0.32, P=0.99 | $Log_2FC$=0.42, P=1.0 | $Log_2FC$=-0.91, P=0.98 |
| IL1B | $Log_2FC$=-0.014, P=0.99 | $Log_2FC$=-0.14, P=0.99 | $Log_2FC$=0.38, P=1.0 | $Log_2FC$=-0.29, P=0.98 |

**Figure S4. Overall survival (OS) and progression-free survival (PFS) by mutated CH gene**

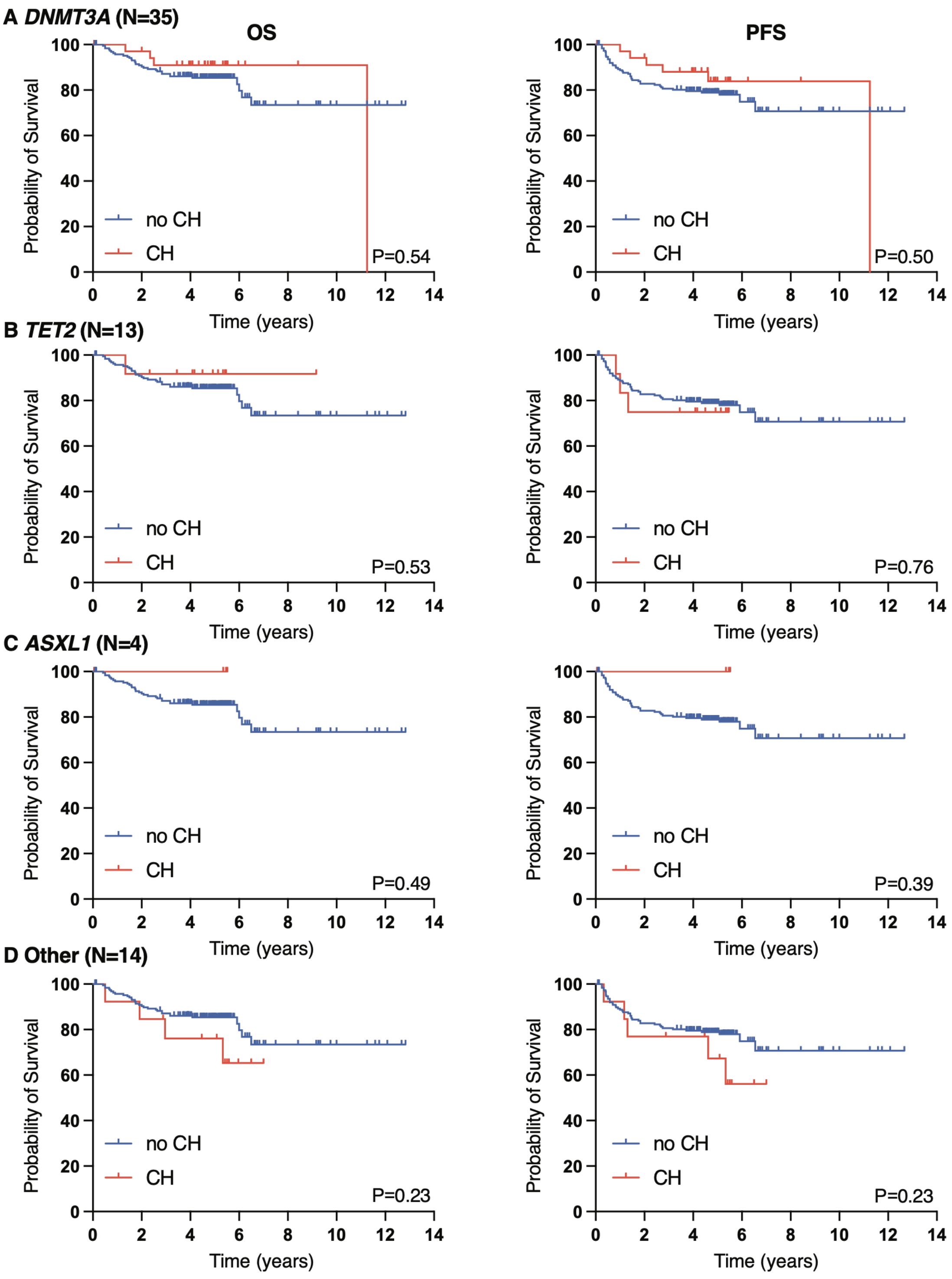

**Figure S5. Multivariable models for OS and PFS in each trial**

**A: NLG-LBC-04 (N=4)**

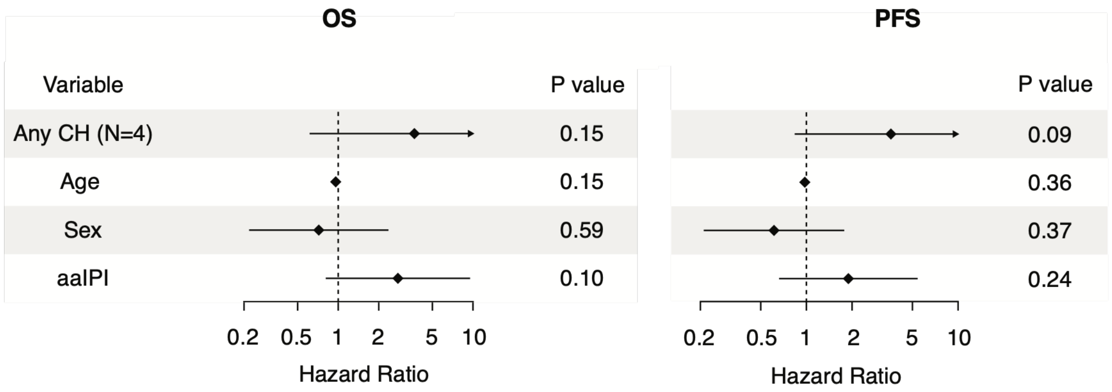


**B: NLG-LBC-05 (N=28)**

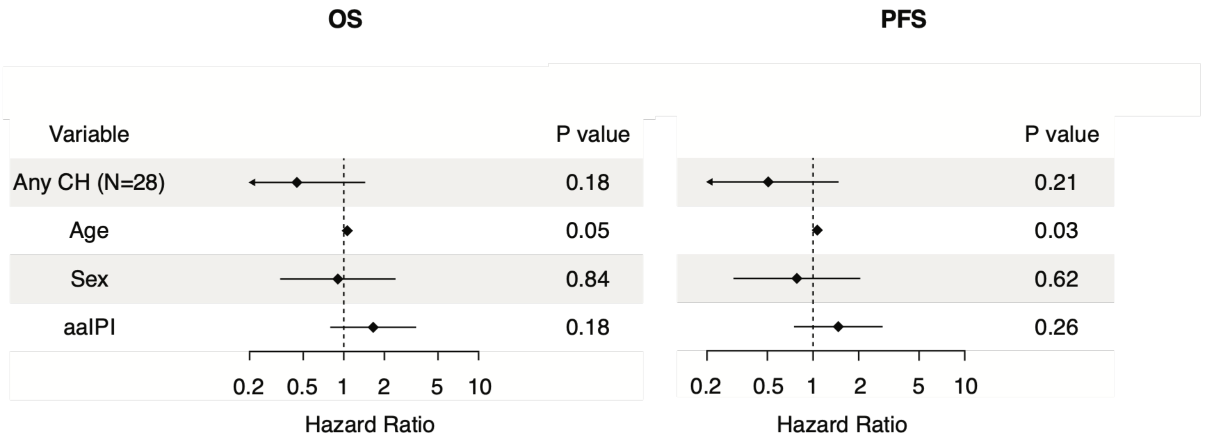


**C: NLG-LBC-06 (N=25)**

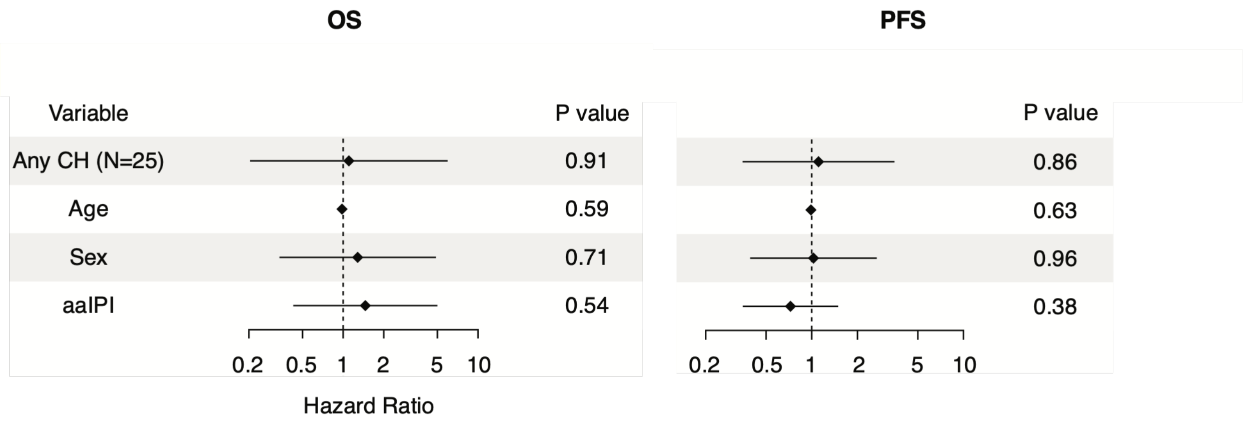

### Figure S6. CH variant allele frequencies in PB, tumor, and cfDNA

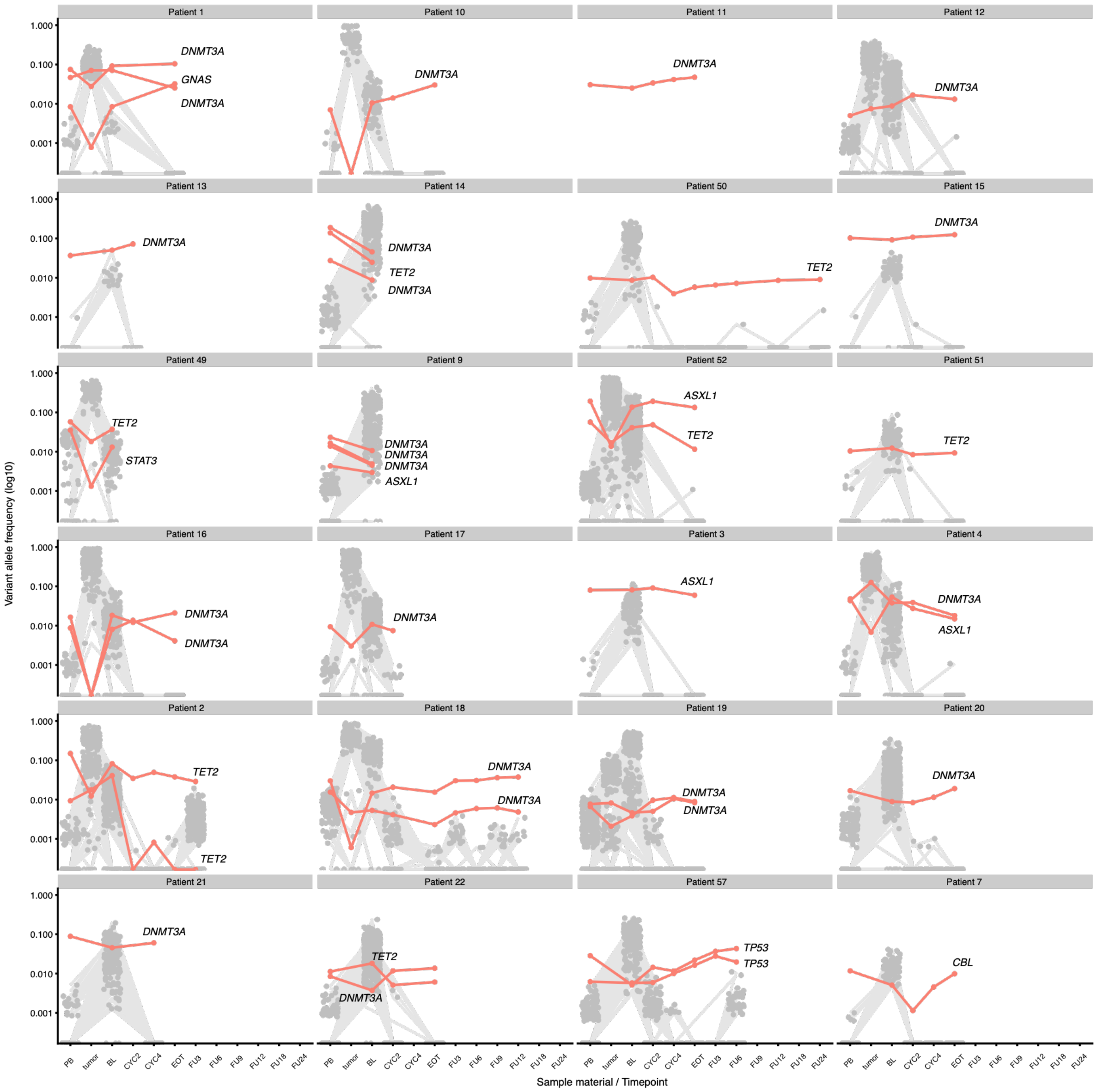

**Figure S7. Relative variant allele frequencies of TET2 and lymphoma variants in cfDNA**

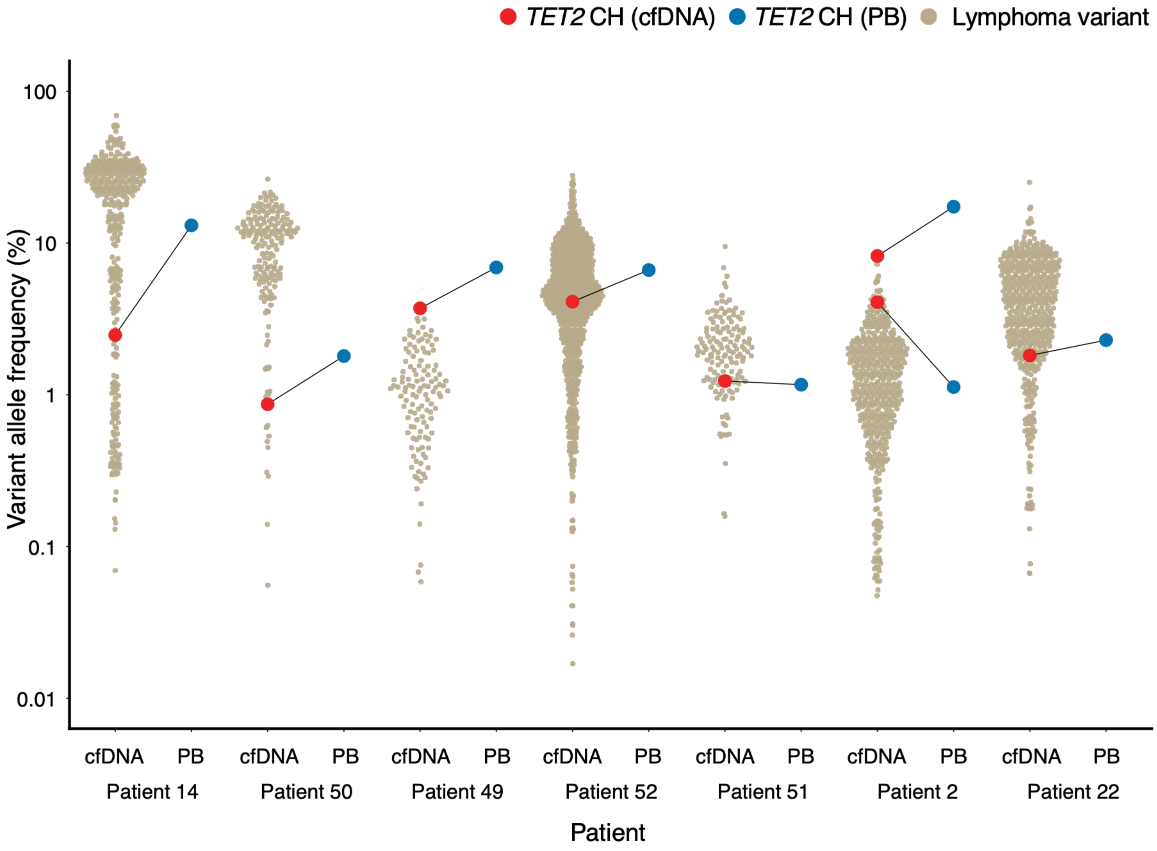
